# A mixed-methods analysis of the implementation of a new community long-COVID service during the 2020 pandemic: learning from practice

**DOI:** 10.1101/2024.10.25.24316101

**Authors:** Stefanie L Williams, Emily Beadle, Paul Williams, Harsha Master, Annalisa Casarin

**Author notes:** Corresponding author: (SW).

## Abstract

**Introduction:** The rapidly increasing prevalence of long-COVID (LC), the multisystem complexity of the condition and high patient symptom burden, necessitated an immediate need to develop new clinics for assessment and management. This article reports on the rapid implementation of a reactive and responsive LC care pathway. We mapped patients’ journey through this pathway, identifying the services that were activated according to prevalent symptoms, and assessed the barriers and facilitators to its implementation and delivery, from the perspective of health care professionals (HCPs) and LC patients using the Theoretical Domains Framework (TDF).

**Methods:** Mixed methods study, including retrospective quantitative cross-sectional analysis of patient data and semi-structured qualitative interviews. One hundred and sixteen patients who attended long-COVID clinic in Hertfordshire, UK, in the first 5 months of its existence, consented for their data to be analysed for the quantitative study. Six HCPs and five patients participated in semi-structured interviews.

**Results:** Patients were referred into the service an average of 5.75 months post initial COVID-19 infection. 82% of patients required onward referral to other HCPs, most commonly pulmonary rehabilitation, chronic fatigue specialists, and the specialist COVID-19 Rehab general practitioner embedded within the service. Patients reported having rehabilitation needs, moderate depression and anxiety, and difficulties performing usual activities of daily living at point of care. The TDF domains most relevant to the implementation of the LC pathway were *beliefs about capabilities, environmental context and resources, knowledge, and reinforcement*.

**Discussion:** Our study provides novel insight into the development of a reactive multidisciplinary care pathway. Key drivers for successful implementation of LC services were identified, such as leadership, multidisciplinary teamwork, transferable skills, and knowledge exchange. Barriers to rapid set up of the service included funding constraints and the rapid evolution of an emergency context.

## Introduction

### Long-COVID

Following recovery from acute COVID-19 infection, some patients subsequently experience a range of persistent and debilitating physical and psychological symptoms such as fatigue, breathlessness, and cognitive impairment. Such symptoms, of which over 200 have been reported, are indicative of a multiphasic, cyclical condition, which has become known by the patient-made term ‘long-COVID’ (LC) [1,2]. The World Health Organization (WHO) defines LC, otherwise known as post-COVID-19 syndrome and Post-Acute Sequelae of CoVID-19 (PASC), as *“the continuation or development of new symptoms 3 months after the initial SARS-CoV-2 infection, with these symptoms lasting for at least 2 months with no other explanation*” [2]. The condition is estimated to affect 1.9 million people in the UK [3] and at least 65 million individuals worldwide[4].

### Long-COVID services

Due to the rapidly increasing prevalence of LC internationally, the need for the development of new care pathways and multidisciplinary clinics to assess and manage symptomatic patients was recognised, and advocated for by patient groups, clinicians, and researchers as early as April 2020 [5,6]. Furthermore, the WHO emphasised the need for multidisciplinary approaches to the assessment and management of LC, which are contextually appropriate and tailored to the specific complexities of the multisystem condition [7]. In 2021, the NHS provided £10 million for the creation of a network of clinics to help assess and treat those in the community exhibiting symptoms of LC, with the target of creating more than 60 centres across England [8]. This was subsequently followed by an additional £90 million investment in specialist LC clinics in England [9]. Prior to the availability of additional resource, community services adapted their care provision in order to absorb the high demand of caring for people with LC. With very limited national guidance or research, there was an immediate need to develop new pathways in response to clinical need.

### Barriers to accessing care for long-COVID

Access to specific LC support can validate patients’ experiences of the condition[10]. However, limited awareness, poor coverage, inconsistency, disconnect between healthcare systems and funding uncertainties have been identified as structural barriers to the provision of specialised integrated care for LC [11]. Healthcare professionals have reported a lack of existing referral pathways, service capacity pressures, lack of medical staff within LC service provision, resource issues, gaps in knowledge and lack of confidence in managing the condition as barriers to the effective implementation of community rehabilitation for LC [12]. However, these findings were reflective of the challenges of delivering adapting existing integrated services, to absorb LC patients, rather than a dedicated LC clinic. Furthermore, existing research examining the influences on the implementation of LC services, from the perspective of those involved in their delivery, is so far limited to two UK regions, and do not refer to frameworks of implementation science [11,12]. Specific barriers identified by groups disproportionately affected by COVID-19, such as ethnic minorities, included mistrust in health professionals and fear [13]. A finding which may explain poorer uptake of multidisciplinary LC clinics by these groups [13,14].

### Implementation science

The implementation, and subsequent ongoing delivery, of a new multidisciplinary healthcare service requires substantial behavioural changes at different levels within the organisation responsible for its inception. The Theoretical Domains Framework (TDF) is a theory-based approach to implementation, which can be used to provide a granular understanding of the psychological capability and reflective motivational processes underpinning behaviour change within this context [15,16]. The TDF[15] consists of 14 theoretical domains relevant to behaviour change: knowledge; skills; memory; attention and decision processes; behavioural regulation; social/professional role and identity; beliefs about capabilities; optimism; beliefs about consequences; intentions; goals; reinforcement; emotion, environmental context and resources and social influences. The TDF has been used extensively across a wide range of healthcare settings to understand barriers and facilitators to the implementation of guidelines and services [17,18]. In relation to COVID-19, the TDF has been used to understand covid-19 disease prevention behaviours, including vaccine uptake [19–22], hand hygiene [19,23] and adherence to guidelines [24]. It has also been applied to the development of an intervention to support doctors’ well-being and resilience during CoVID-19 [25]. However, to our knowledge this is the first study to apply the TDF within the context of the implementation of a LC service to understand the barriers and facilitators to the effective implementation of such a service.

### Aims of the study

The present study reports on the rapid implementation of a reactive and responsive, jointly specialist COVID-19 Rehab General Practitioner (GP) and Applied Health Professional (AHP)-led multidisciplinary pathway in a community setting, developed to help manage demands associated with the long-term complications of CoVID-19. The pathway was first set up in August 2020, prior to the National Institute for clinical Excellence (NICE) publication of case definition in November 2020 and clinical guidance in December 2020 [26]. Referrals into the service came from both primary and secondary care. In addition to examining the demographic and symptom characteristics of the patients accessing the pathway and mapping their journey through it, which can provide comparison with other long-COVID pathways [14], we also aimed to explore the experiences of service users and providers. In particular, we sought to assess the barriers and facilitators to the implementation of the clinic from the perspective of both LC clinic HCPs, and LC patients receiving care via, the pathway.

## Materials and Methods

### Study design and setting

The study took place within a Hertfordshire LC service, which began in August 2020. Covering a population of 600,000 people in East and North Hertfordshire, the multi-disciplinary team (MDT) initiated and developed a new COVID-19 pathway in response to patient need and demand underpinned by an integrated approach to care i.e. to provide coordinated, holistic care involving both medical assessment and rehabilitation. A COVID-19 rehabilitation register was developed to identify and map patients in need of support and to track patient numbers and outcomes across the NHS trust. An Allied Health Professional COVID-19 coordinator was recruited to facilitate the coordination and triage of these patients across pathways and systems from August 2020. The pathway was an entirely virtual community-based clinic jointly led by a Specialist COVID-19 Rehab General Practitioner (GP) and Occupational Therapist, with a large MDT of allied health professionals. These included Physiotherapists, Speech and Language Therapists, Dietitians, Pulmonary Rehab, Chronic Fatigue specialists, and a Clinical Psychologist. Complex patients were reviewed by the clinic’s GP and referred on to secondary care specialists as needed. The team also worked closely with colleagues in the acute hospitals, social care, and the voluntary sector.

A mixed-methods study design was used for this evaluation. The quantitative component consisted of a retrospective cross-sectional analysis of data collected from patients admitted to the LC service between 1st August 2020 and 31st December 2020. The qualitative component consisted of semi-structured interviews with 1) a subsample of these patients, and 2) HCPs working within the service during the same time period.

### Participants and Data collection procedures

Patients and HCPs were eligible to participate if they were referred to or worked within the service between the dates specified above. There were no limitations on eligibility to take part according to HCP type. Where possible, purposive sampling was used for participant selection for the qualitative interviews to ensure inclusion of participants with diverse characteristics. The recruitment period for this study was between 14^th^ April 2022 and 31^st^ July 2023.

Quantitative data were collected by staff within the clinic (HM) during the day-to-day running of the service and recorded (EB) using an electronic data collection form developed specifically for this study (S1 Table). Upon entering the clinic, a clinical assessment was performed, a post COVID-19 rehabilitation form filled in and patients were asked to complete questionnaires including the measures detailed below. Of 218 patients referred into the clinic, 116 provided consent for their data to be analysed for research purposes. Of those, five patients agreed to participate in a semi-structured interview. Patients were approached to participate in the research via email and/or telephone. In total, six clinicians were invited to participate, and all agreed to participate in a semi-structured interview. All participants were provided with information sheets and provided written consent by signing consent forms prior to participation in the interview.

All qualitative interviews were conducted by the same male researcher (PW), an Academic Research Fellow (PhD) at the time of the interview. PW knew all clinicians because of working within the clinic in a research capacity, PW did not have a relationship with any of the patients prior to study commencement. Participants were provided with the researcher’s name and contact details in the information sheets, as well as reasons for doing the research. No other detail related to the interviewer were provided. Interviews took place online and were recorded. Only the interviewer and participants were present during the interviews. Patient and HCP interviews lasted approximately 60 minutes. Field notes were taken during the interview. No repeat interviews were undertaken. After interviewing five patients and six clinicians, the research team were satisfied that data saturation had been reached, recruitment was then ceased. Quantitative data were accessed on 14^th^ April 2022. Quantitative data were directly accessed by EB prior to analysis to identify eligible cases, EB therefore had access to information that could identify individual participants at this stage. However, quantitative data were anonymised and encrypted by the NHS Trust Information Manager prior to analysis. Interview recordings and verbatim transcript were stored securely for analysis by the authors (AC, EB, PW, SW). Transcripts were not returned to participants for comment and/or correction.

### Measures

Quantitative measures included demographic variables (i.e. age, gender, ethnicity, smoking status, BMI, deprivation status using postcode); Pre-existing comorbidities; Disability (physical, learning). Referral data included date of referral, primary reason for referral, source of referral, discharge data, intervention (rejected for referral or discharged), referrals on to individual services, and date of onwards referral. COVID-19-related test/ history data included i) hospitalisation, ii) current/past treatment, iii) onset, iv) test results, v) initial symptoms, and v) ongoing problems.

In addition, the following measures were used: Yorkshire Rehabilitation Screening (C19-YRS)[27,28]; frailty scale; Malnutrition universal screening; UCLH loneliness assessment; PHQ-9 [29]; GAD-7 [30]; Patient functional scale and self-management plan; EuroQol five-dimension health questionnaire (EQ5D) [31]. The PHQ-9[29] and GAD-7 [30] were only completed when deemed necessary by LC clinic staff, and the C19-YRS[27,28] measure was developed during the COVID-19 pandemic. EQ5D [31] data were not initially collected upon clinic set up. Thus, completion rates for these measures were lower than for the other included measures.

Qualitative data were collected through semi-structured interviews. Patient interview schedules developed by the authors (AC, EB) focused on; patients overall experience of the LC clinic, clarity of their assessment and treatment pathway through the clinic, interactions with staff/clinicians, clarity of communication with clinicians/staff, satisfaction with received care, potential improvements of care and pathway, preference for virtual or in-person services. HCP interview schedules included questions on their experiences of setting up a reactive LC service, of working in a multi-disciplinary team, the organisational structure of the service, issues experienced delivering care in time of a pandemic, virtual services, interactions with patients, interactions with other staff/clinicians, adaptations to service based on patient needs/wants. Topic guides for patient and HCP interviews are available as supplementary material (S2 File).

## Data analysis

### Quantitative data analysis

Patient demographic variables are reported using frequencies and/or percentages for categorical variables and means ± SD for continuous variables. Descriptive data on service utilisation and referral pathways are reported using frequencies and/or percentages, including i) where patients were referred from, ii) primary reason for being referred into to the service, iii) which services were used by which patients, iv) number of services accessed by each patient, v) referrals on to other services. Pearson’s chi-square/Fisher’s exact was used to explore the relationships between demographic and service-related variables (e.g. sex and primary reason for referral). Data were analysed in SPSS (version 28) by EB.

### Qualitative data analysis

Anonymised interview data were coded using NVivo (version 12). Data were analysed using thematic analysis [32]. Inductive open coding using a line-by-line process was first used to analyse the transcripts. To ensure integrity of the analysis process, two coders (PW, AC) independently coded the first two interviews. Following this, the rest of the interviews were analysed by three coders (PW, AC, EB).

Quotes were deductively allocated to one of 14-domains specified within the TDF [15] using a table developed for the current study. To prevent omitting important data, subdomains were added where appropriate. Discrepancies in allocating quotes to a relevant TDF domains were resolved by discussion. A framework matrix was developed to reduce and organise the data into themes, cases, and sets, and connections between categories were mapped to explore relationships and/or causality. Data triangulation was used to gain a greater understanding of multiple perspectives; similarities and differences between domains related to patients’ and healthcare professionals’ views were identified. Finally, codes related to barriers and/or facilitators were extracted and used to create a list (table 3). Participants did not provide feedback on the findings.

### Ethics approval

Ethical approval for the study was obtained through Health Research Authority (HRA) and Health and Care Research Wales (HCRW) Research Ethics Committee (REC reference: 21/PR/0987). All participants provided informed consent prior to participating in the study. University of Hertfordshire and institutional ethics guidelines were followed.

### Patient and public involvement

The project was designed to capture the efforts of the community care professionals to meet patients’ expectations for LC assessment and management. We believe the aims of the study reflects patients’ priorities in research. Exploring their experiences in accessing care and navigating through a newly developed pathway was discussed with members of the public who attended clinic appointments. A person responded welcoming the study and confirming the importance of research on all things related to LC. We also sought involvement from HCPs involved in delivery of the LC clinic who contributed to shaping the aims and study plan. We will seek advice from study participants on the best methods to reach the public with the study findings.

## Results

Between August and December 2020, 218 patients were referred to the clinic, of which 116 gave consent to take part in the current study. All patients were diagnosed with LC, had suspected LC, or had COVID-19 associated rehabilitation needs.

### Patient characteristics

Patients attending the service (n=116) were aged between 19 and 83 years (Mean = 50.68, SD = 14.40) and the majority were female (70.7%). Most were classified as living in non-deprived conditions (98.3%) and were White British (76.7%). The majority were non-smokers (75%) or ex-smokers (19%), only 3.4% were current smokers. The mean number of comorbidities patients reported ranged between 0-6 (mean= 1.95). The most common were chronic pain/fatigue (21.55%), unspecified respiratory illness (20.68%), heart disease (18.97%), asthma (16.37%) and hypertension (11.2%). See Table 1.

**Table 1.** Characteristics of patients entering the LC clinic.

|  |  | N | (age range) and<br>% of study population | Mean | SD |
| --- | --- | --- | --- | --- | --- |
| Age |  | 116 | (19-83yo) | 50.68 | 14.40 |
| Sex | Male | 34 | 29.31% |  |  |
|  | Female | 82 | 70.69% |  |  |
| Deprivation | Deprived | 2 | 1.72% |  |  |
|  | Non-Deprived | 114 | 98.28% |  |  |
| Ethnicity | White British | 89 | 76.72% |  |  |
|  | White Irish | 2 | 1.72% |  |  |
|  | White Other | 5 | 4.31% |  |  |
|  | Indian or Indian British | 3 | 2.59% |  |  |
|  | Caribbean | 1 | 0.86% |  |  |
|  | British or Mixed British | 10 | 8.62% |  |  |
|  | Mixed Other | 2 | 1.72% |  |  |
|  | No Data | 4 | 3.45% |  |  |
| Smoking-Status | Smoker | 4 | 3.45% |  |  |
|  | Ex-smoker | 22 | 18.97% |  |  |
|  | Non-smoker | 87 | 75% |  |  |
|  | Info refused/not known | 3 | 2.59% |  |  |
| Co-morbidities | Unspecified respiratory illness | 24 | 20.68% |  |  |
|  | Asthma | 19 | 16.37% |  |  |
|  | COPD | 5 | 4.31% |  |  |
|  | Diabetes | 8 | 6.89% |  |  |
|  | Hypertension | 13 | 11.20% |  |  |
|  | Heart Disease | 22 | 18.97% |  |  |
|  | Hyper/Hypo-thyroidism | 7 | 6.03% |  |  |
|  | Cancer | 4 | 3.45% |  |  |
|  | Skin condition | 8 | 6.89% |  |  |
|  | Migraines/chronic headache | 5 | 4.31% |  |  |
|  | Sleep condition | 3 | 2.59% |  |  |
|  | High BMI | 7 | 6.03% |  |  |
|  | Mobility | 6 | 5.17% |  |  |
|  | Chronic pain/fatigue | 25 | 21.55% |  |  |
|  | Gastrointestinal Disease | 11 | 9.48% |  |  |
|  | Other | 24 | 20.68% |  |  |

Patients within the LC service reported having ongoing rehabilitation needs (C19-YRS mean 7.99, SD= 6.31). Patients also reported having moderate depression (mean= 13.21, SD= 7.22) and moderate anxiety (mean= 9.39, SD= 6.76) associated with onset of COVID-19 and reported difficulties with performing usual activities of daily living (mean= 1.95, SD= 1.01).

There were no significant differences shown between month of referral into the service and C19-YRS, PHQ-9, and GAD-7 outcomes. However, there was an increase in the average score of the C19-YRS in November 2020. Patients reporting gastro-intestinal comorbidities reported significantly lower scores on EQ5D self-care measured at access to the service (mean = 1.0), versus those without (mean = 1.30), t (98) = 4.10, p<.001. No other significant differences observed regarding comorbidity type or frequency and EQ5D.

### Referrals into the service

There were between 15 and 30 patients (mean=23.2) referred into the service per month, during the five-month study period. The number of patients referred into the service during August, September, October, November and December was 30, 20, 27, 24 and 15, respectively.

Patients were admitted to the LC clinic on average 5.75 months post initial-infection/symptoms of COVID-19. The majority of patients were admitted 7 months post-COVID (N=28), reflecting an absence of LC clinics prior to August 2020, e.g., a person being affected by the infection in the early part of the year (Jan-April) and being admitted to the clinic between Aug and Nov. One patient was admitted in the same month as they experienced COVID symptoms.

The majority of patients were referred to the service by their General Practitioner (GP: 72.4%). Initial symptoms of COVID-19 were treated at home by 63.8% of patients and the majority did not have a test (41.4%), or were too early for testing (18.1%), to confirm their COVID-19 status. In total, 33.6% had a positive test result confirmed on referral to the service. Most patients were referred to the service for respiratory conditions (50.9%) or chronic fatigue issues (18.1%). The journey of patients through the service pathway can be found in S3 Figure.

### COVID and Long-COVID symptoms of patients referred into the service

Temperature changes (increased, reduced or both; n=59, 50.86%), shortness of breath (SOB; n=76, 65.5%), cough (n=59, 50.86%), and fatigue (N=51, 43.97%) were the most reported symptoms for initial acute COVID infection. There was a significant association between shortness of breath (SOB) at initial infection and smoking status, with non-smokers (never smoked) more likely to report SOB than current smokers and ex-smokers (p= 0.022). See Fig 1.

**Fig 1.**
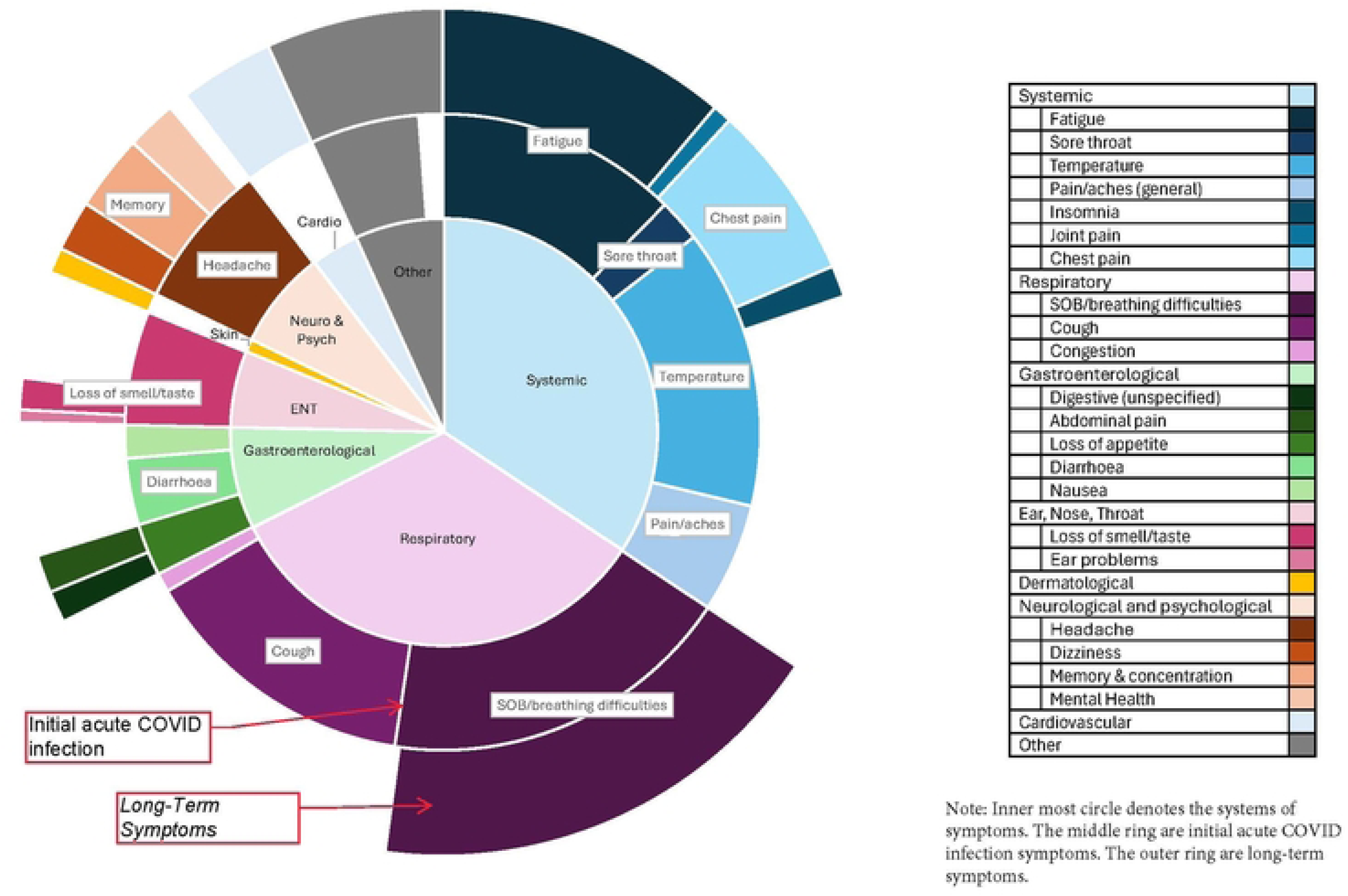
COVID and long-COVID symptoms of patients referred into the service.

Breathing difficulties, primarily continued shortness of breath (n=74,63.79%), and fatigue (n=46, 39.66%) were the most reported LC symptoms at admittance to the service.

Fatigue and breathing difficulties were present in both primary COVID infection and LC, while cardiac (n=16, 13.79%), concentration (n=12, 10.34%) and mental health issues (n=8, 6.89%) started only after the primary infection.

There was a significant difference in primary COVID infection symptoms between patients who were hospitalised and those that were not hospitalised (i.e. homecare, A&E access but not admitted, self-referral). People who were treated at home or were admitted to A&E and subsequently discharged were more likely to report pain/aches (p = .011), headache (p = .043), changes in smell/taste (p = .008) and fatigue (p = .002). Patients referred into the service from primary care were significantly more likely to report pain/aches (p=.039) and changes in smell/taste (p=.040) at primary infection than patients referred to the service from secondary care.

### Referrals out of the service

There was an average of 1.59 unique referrals out of the service on to other services and health professionals, per patient (range 0 to 5). In total, 82% of patients had an onward specialist referral. The majority of patients required referral on to pulmonary rehabilitation (n= 65, 56.03%). The other services patients were referred to were; Specialist COVID Rehab General Practitioner (n=44, 37.93%), Community Chronic Fatigue Specialist (n=21, 18.1%), Occupational Health (n=19, 16.37%), Psychologist (n=10, 8.62%), Mental Health Team (n=7, 6.03%), Community Nurse (n=5, 4.31%), Dietitian (n=4, 3.45%), Respiratory Nurse(n=3, 2.59%), Cardiology (n=4, 3.45%), Respiratory Physician (n=3, 2.59%), Physiotherapy (n=2, 1.72%), and Neurology (n=1, 0.86%).

Patients who were hospitalised were more likely be referred to further specialist advice e.g. cardiology (dietitian, psychology), whilst patients who were not hospitalised were more likely to be referred to the specialist COVID-19 Rehab GP and pulmonary rehabilitation for further care (Fig 2).

**Fig 2.**
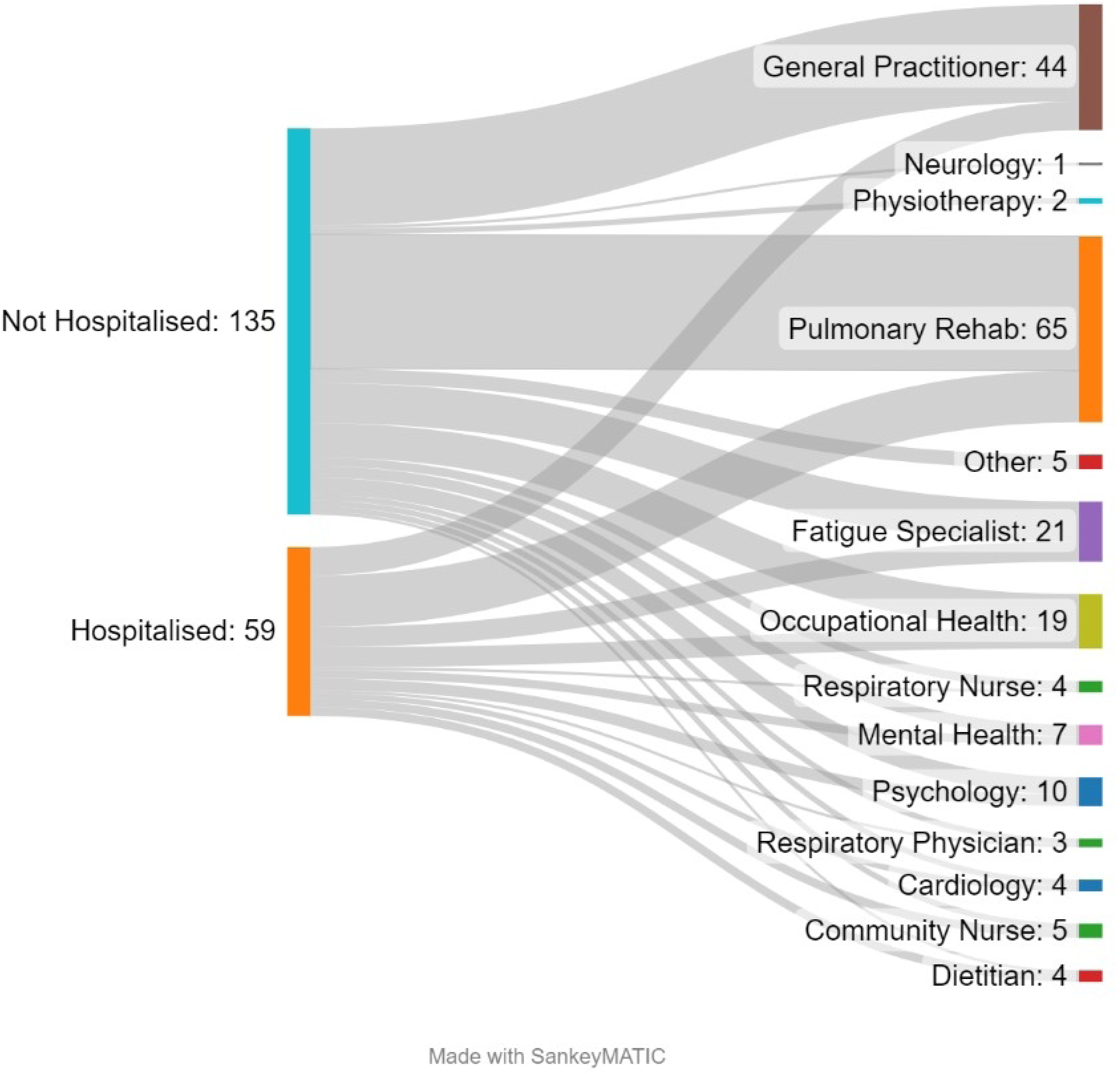
Referrals out to other services.

Patients with higher EQ5D pain scores had significantly more outward referrals than those with less pain (F(5, 104) = 2.64, p = .027). Patients with anxiety and/or depression symptoms, measured using EQ5D, had significantly more unique outward referrals than patients without anxiety and/or depression (F(5, 104) = 2.58, p = .030). There was a significant association between gender and outward primary care referrals (e.g., those referred to the specialist COVID-19 Rehab GP, community nursing, community chronic fatigue syndrome specialist, and pulmonary rehab). Females were significantly more likely to be referred to the specialist COVID-19 Rehab GP than males [X2(1) = 5.88, p = .020]. There were no association between gender and outward referrals to mental health specialists (p = .78), allied health professionals services (p = .30), or secondary care (p = .40).

### Discharge from long COVID service

In total, 90 patients (77.59%) were discharged from the service. Of these, 62% (n=56) had significantly improved symptoms or were asymptomatic, whilst 38% (n-34) were still symptomatic. One patient was discharged but requested to be re-admitted to the service. Twenty-five patients remained within the service. There was a significant association between long-term (LC) associated self-reported fatigue and discharge status (p = .002), with those without fatigue significantly more likely to have significantly improved symptoms of LC and were discharged, while those with fatigue were more likely to remain in the service.

### Qualitative data

In total, eleven qualitative interviews were conducted, with five patients and six health care professionals (HCPs); two doctors, one physiotherapist, two occupational therapists, one coordinator). TDF [15] domains indicated as most important and relevant to the implementation of a new service to address LC patient needs, based on our analysis of patient and HCP interview data, are presented in table 2. The number of participants who mentioned, and the number of references related to, a specific domain is also presented. Where appropriate, sub-domains were added to further capture important features of some TDF domains.

**Table 2.**
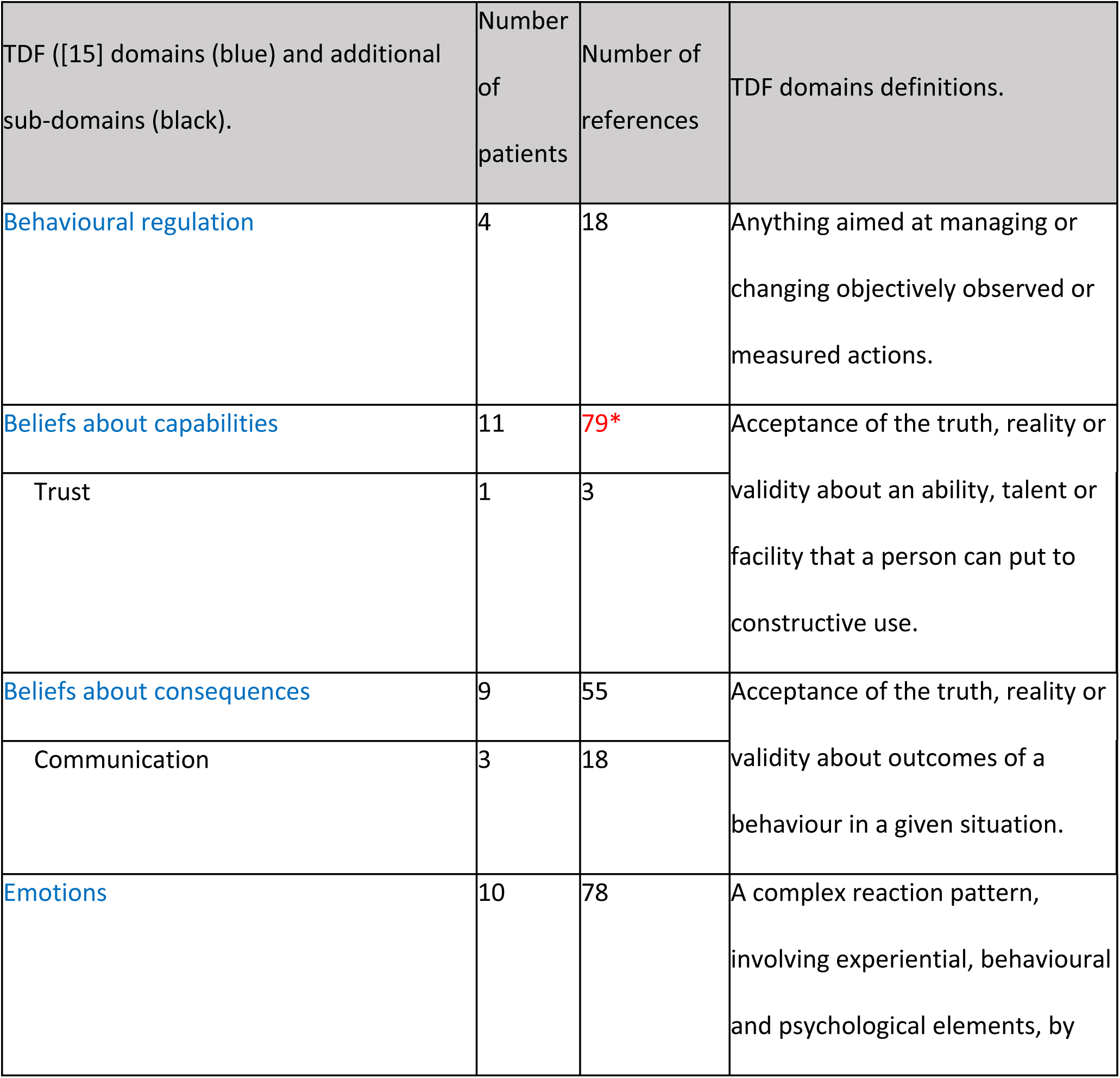
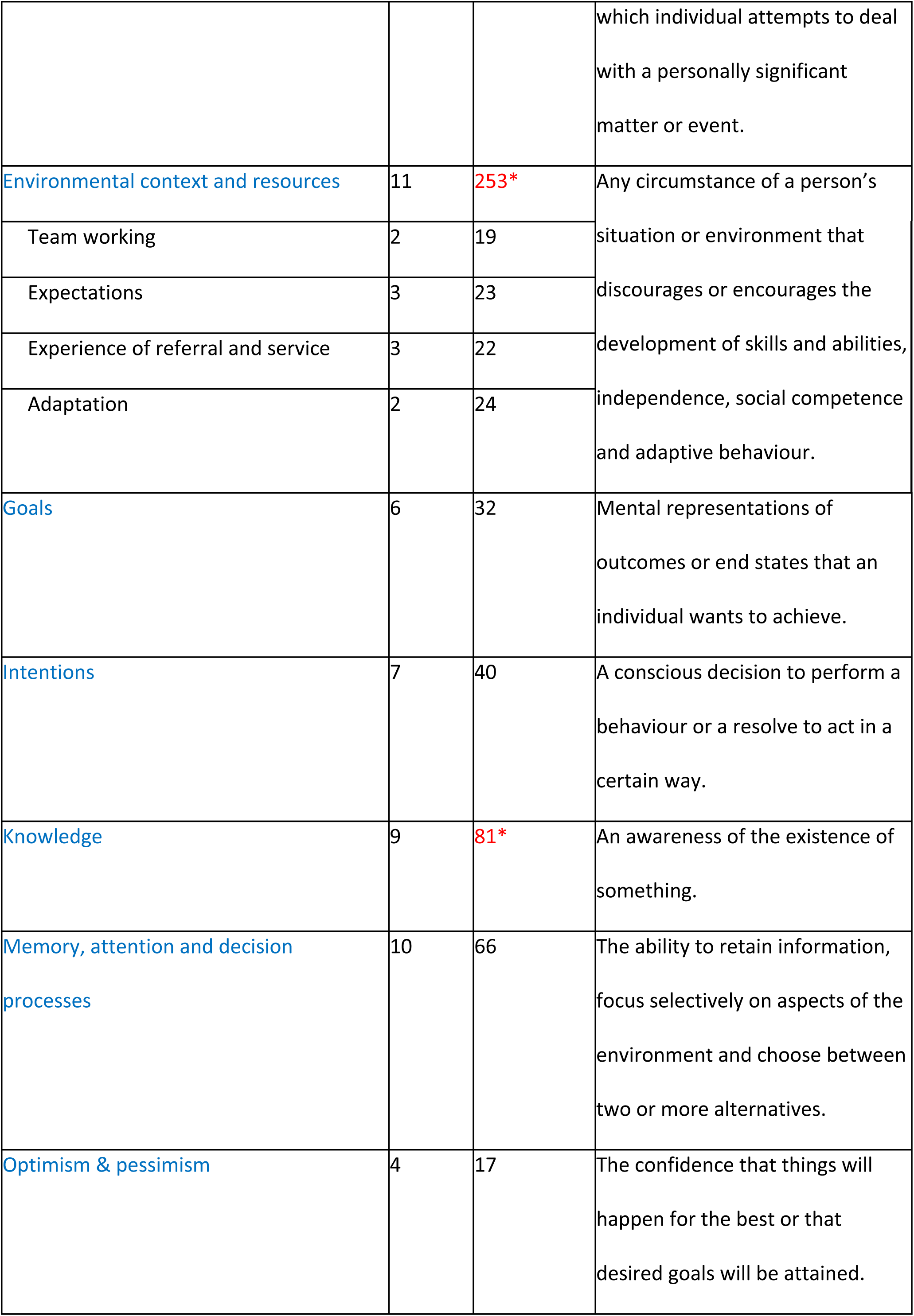
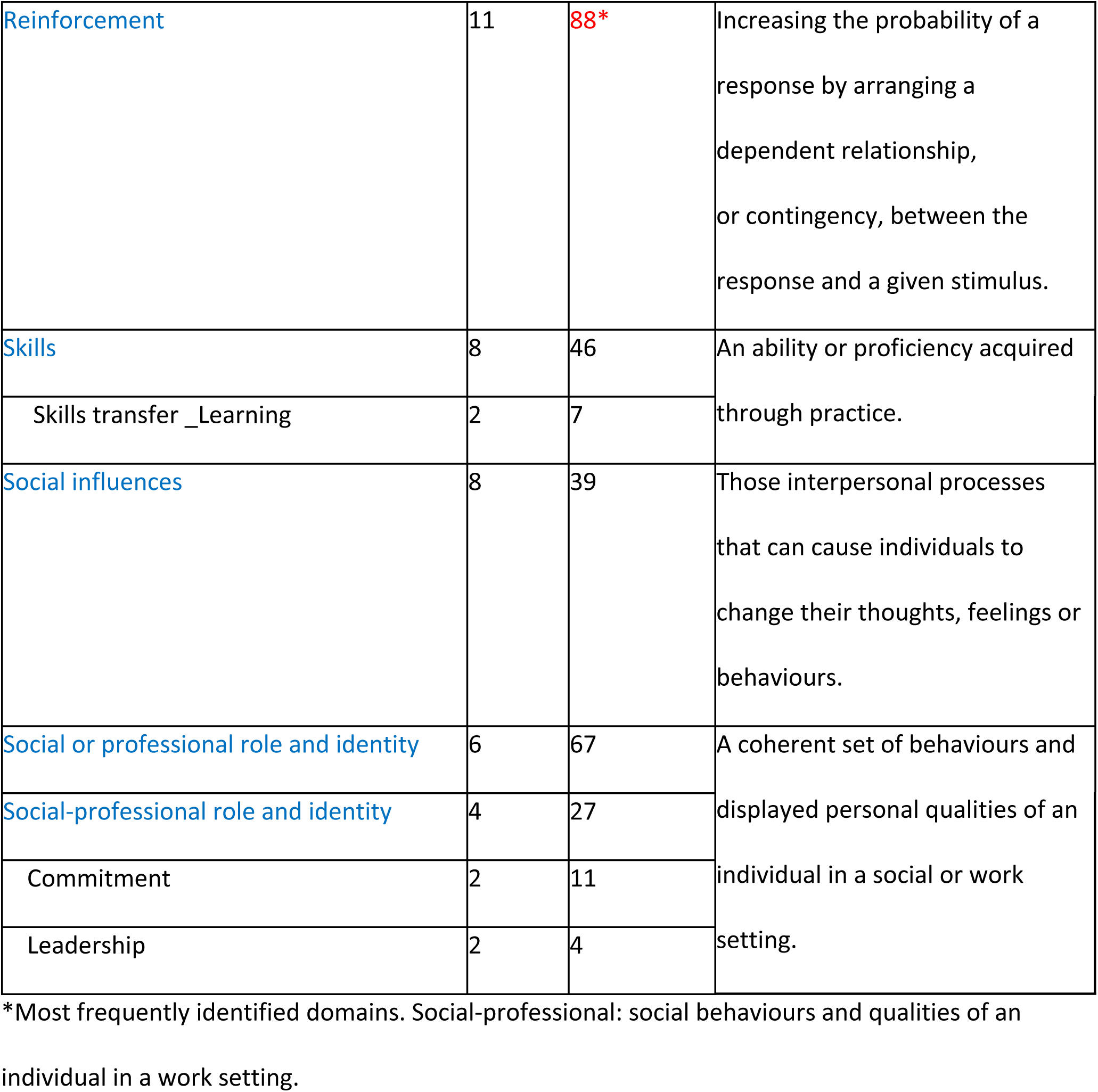
TDF [15] domains and subdomains across patients and HCPs.

| TDF ([15] domains (blue) and additional sub-domains (black). | Number of patients | Number of references | TDF domains definitions. |
| --- | --- | --- | --- |
| Behavioural regulation | 4 | 18 | Anything aimed at managing or changing objectively observed or measured actions. |
| Beliefs about capabilities | 11 | 79* | Acceptance of the truth, reality or validity about an ability, talent or facility that a person can put to constructive use. |
| Trust | 1 | 3 |  |
| Beliefs about consequences | 9 | 55 | Acceptance of the truth, reality or validity about outcomes of a behaviour in a given situation. |
| Communication | 3 | 18 |  |
| Emotions | 10 | 78 | A complex reaction pattern, involving experiential, behavioural and psychological elements, by which individual attempts to deal with a personally significant matter or event. |
| Environmental context and resources | 11 | 253* | Any circumstance of a person's situation or environment that discourages or encourages the development of skills and abilities, independence, social competence and adaptive behaviour. |
| Team working | 2 | 19 |  |
| Expectations | 3 | 23 |  |
| Experience of referral and service | 3 | 22 |  |
| Adaptation | 2 | 24 |  |
| Goals | 6 | 32 | Mental representations of outcomes or end states that an individual wants to achieve. |
| Intentions | 7 | 40 | A conscious decision to perform a behaviour or a resolve to act in a certain way. |
| Knowledge | 9 | 81* | An awareness of the existence of something. |
| Memory, attention and decision processes | 10 | 66 | The ability to retain information, focus selectively on aspects of the environment and choose between two or more alternatives. |
| Optimism & pessimism | 4 | 17 | The confidence that things will happen for the best or that desired goals will be attained. |
| Reinforcement | 11 | 88* | Increasing the probability of a response by arranging a dependent relationship, or contingency, between the response and a given stimulus. |
| Skills | 8 | 46 | An ability or proficiency acquired through practice. |
| Skills transfer _ Learning | 2 | 7 |  |
| Social influences | 8 | 39 | Those interpersonal processes that can cause individuals to change their thoughts, feelings or behaviours. |
| Social or professional role and identity | 6 | 67 | A coherent set of behaviours and displayed personal qualities of an individual in a social or work setting. |
| Social-professional role and identity | 4 | 27 |  |
| Commitment | 2 | 11 |  |
| Leadership | 2 | 4 |  |

\*Most frequently identified domains. Social-professional: social behaviours and qualities of an individual in a work setting.

**Table 3.**
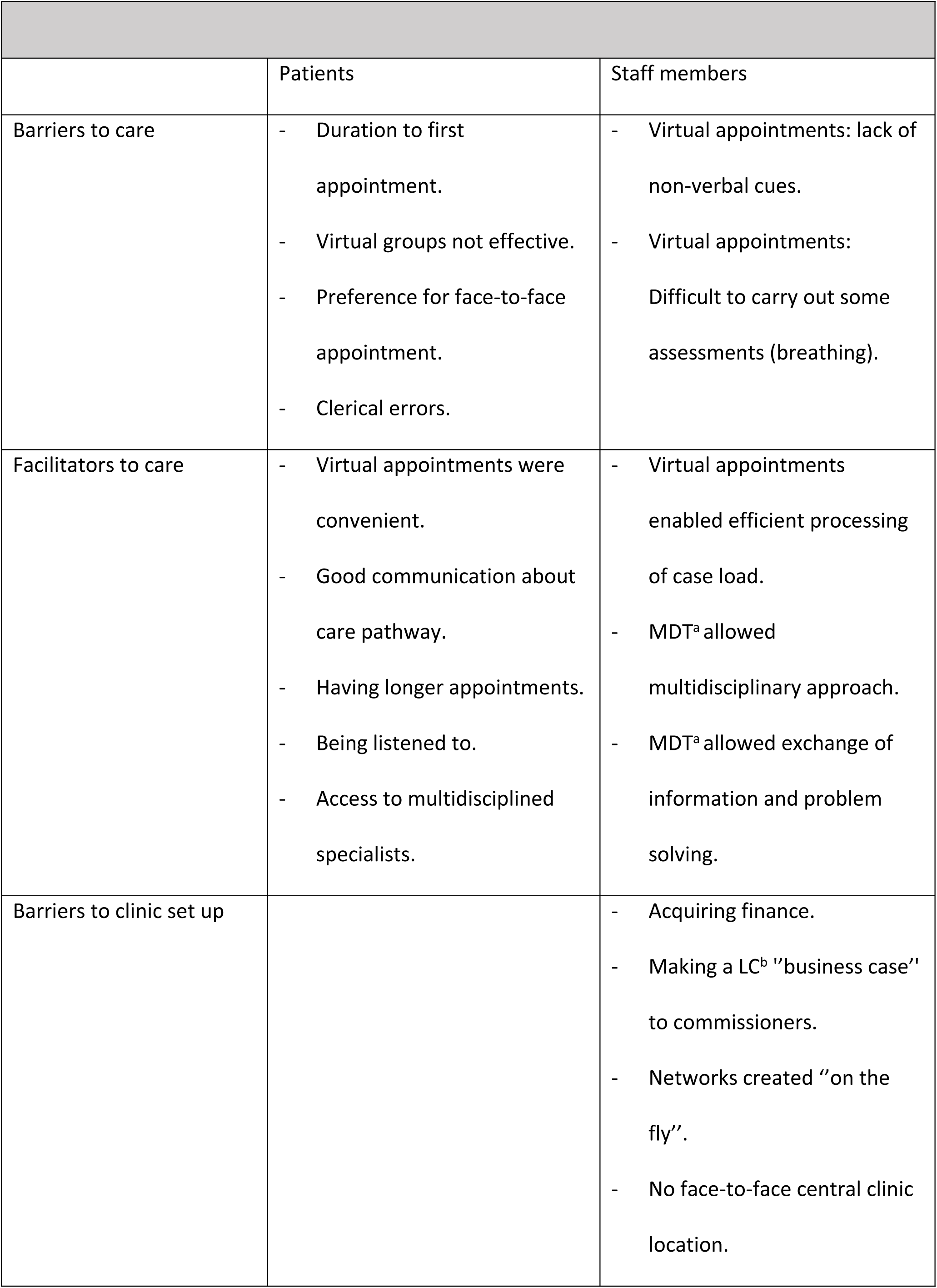
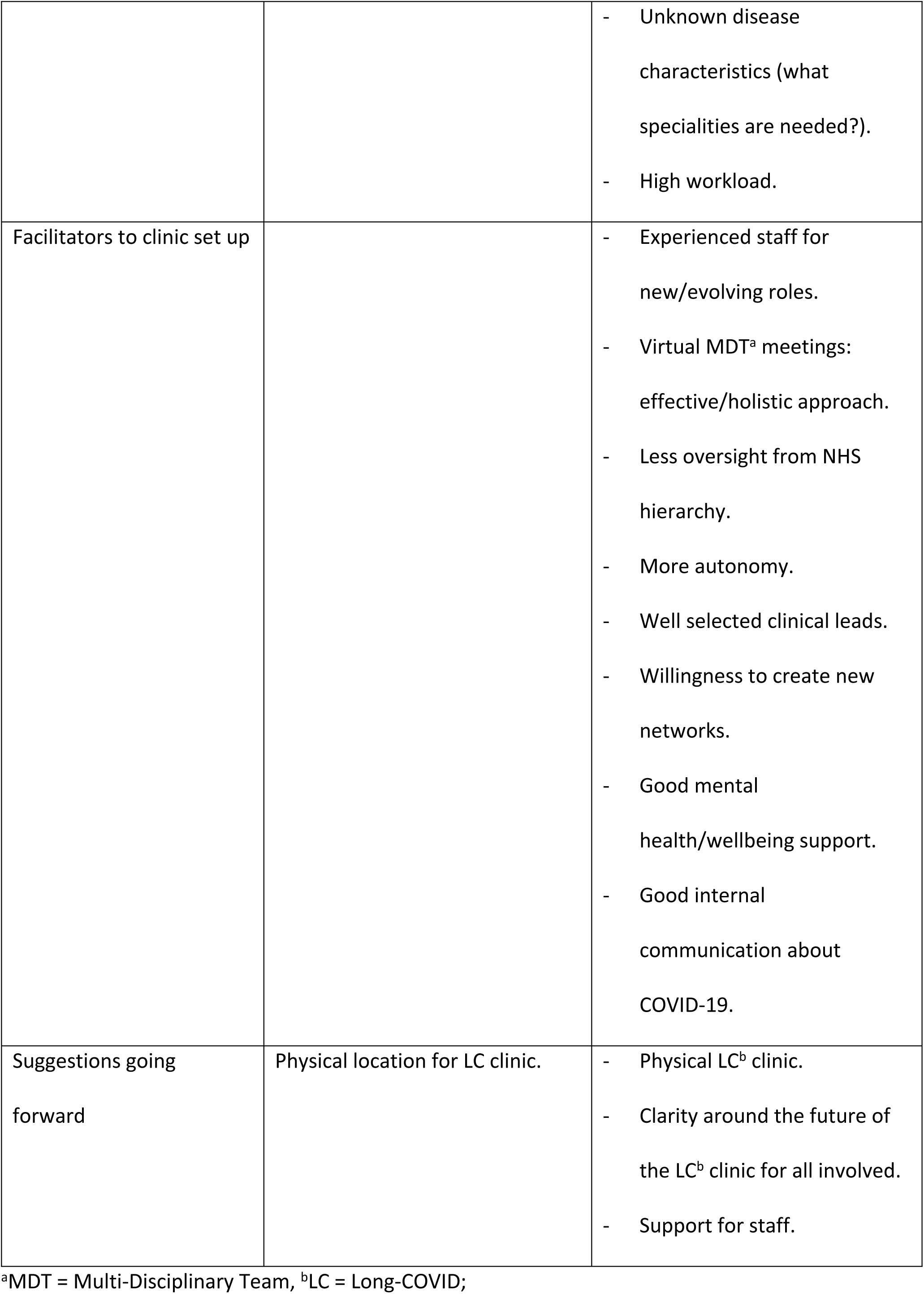
Barriers and facilitators to care and care delivery according to patients and professionals.

Due to the extensive list of domains and sub-themes, the most frequently referenced domains across both patients and HCPs (*beliefs about capabilities, environmental context and resources, knowledge and reinforcement*) are presented in coding trees (S4 Text). A separate coding tree is presented for patients and HCPs. The other domains were also referenced by study participants, and it was found they roughly overlapped with the four dominant domains.

### Themes

The most dominant TDF domains for both patients and HCPs were *beliefs about capabilities* (of staff), *environmental context and resources* (for creating a new care pathway), *knowledge* (of new conditions, its consequences, and new pathway), and *reinforcement* (when a positive experience creates reassurance and confidence on the system).

#### Beliefs about capabilities

Patients stated that the provision of a specific LC service made them feel confident they were well looked after:

> *‘’ I was thrilled to be honest because I just felt quite desperate at the time.’’ P003 ‘’*
>
> *…you felt confident in the process of diagnosis and then subsequent treatment.’’ P008*

Patients acknowledged the new condition was a challenge, but the new service was important, and its existence was valuable:

> *‘’ it was a relief to know that, actually, I wasn’t just gonna be forgotten.’’* P005
>
> *‘’There wasn’t another route I could take. Speak to somebody else about it*.’’ P009

For staff members, the new condition represented a challenge because of the uncertainty of what they were dealing with and the new pathway to be established:

> *‘’ …had to create a pathway where patients could get referred in you know having… making sure that we haven’t missed anything else.*’’ S001

> *‘’ …there was the first scoping meetings, I kind of said, because I’d come from a very systems approach in my previous jobs… I said: you’re gonna have to… have lots of systems involved with this patient cohort because we don’t know what’s needed*. … *I’d kind of have that confidence of working in pathways of care, not in an organizational silo, you know which a lot of people were still very, you know, still office.*’’ S002

> *‘’… quite quickly you realize that you… you didn’t feel like you were as effective in your care. You know, so much of the things that we can ascertain about a patient, we obviously do visually*… *when we started to see post COVID patients face to face. I remember being quite surprised because over the phone they would have a number of symptoms that would be concerning and then they would come into clinic and you would be reassured that actually well, no, you don’t look too bad.’’ S004*

> *‘’ …there was a bit of apprehension because it’s a change and it’s like how is this gonna work from you know various team members but actually within the space of perhaps about six weeks then that waiting list just shrunk then*.’’ S005

> *‘’So, I think I was fairly confident that we could provide a good standard of care in terms of we could do a full assessment on them, and we could direct them to appropriate resources.*’’ S006

#### Environmental context and resources

Patients reflected on the importance of providing the necessary time for staff to assess their conditions and options:

> *‘’ It wasn’t like just a quick 10-minute conversation. Not like I had a bit longer, you know, to properly go into things.’’ P003*

> *‘’ there’s this thing called long COVID. That’s some people are still suffering with effects afterwards and I don’t think those people would if they haven’t been in hospital. I think they’ve missed. They’re missed out.’’ P009*

Capacity of the service was referred to as a barrier to access, and as dependent on resources during a pandemic, when many people needed it:

> *‘’I feel like I was quite lucky. Actually, when I got referred when I sort of been in touch with my GP, I seemed to manage to get referred over quite relatively quickly. I know a lot of people subsequently had quite long waits just because of the sheer numbers.’’ P003*

Experience and the attitude of the professionals was also mentioned as an important feature:

> *‘’ And the lady was just… I seem to remember, being easy to talk to, but she was also… a no-nonsense lady and I liked that, she was very sort of direct and to the point and she knew what she was doing and you felt that this was somebody… in amongst all of this craziness and madness, here was somebody that actually knows… what’s going on because she’d obviously, I’m guessing, drawn on her previous backgrounds.’’ P008*

The pandemic restrictions didn’t affect the quality of the service. Moreover, according to the HCP interviewed, team working excelled. Appreciation was expressed towards leadership; the leaders were firm but approachable.

> *‘’We haven’t had anyone that’s come back and moaned that we haven’t seen them face to face and… and you know, again, when I was a bit confused or if I was stuck or if I was, I would do a video call. And then sometimes I would get like, say, calling rehab… I’d be like, can you see them face to face and let me know what you think.’’ S001*

> *‘’A lot of work from a therapeutic point of view is making sure that people access the right management.’’ S003*

> *‘’ half a day, a week that they had funding for an OT, so very limited when you consider sort of what the need was and what the scope was… So, I was drafted into sort of say, can you take this on because literally there was nobody else.’’ S005*

> *‘’ …had to think about everything in terms of how well we’re gonna gather the data? What was the new system? How the template gonna look like for this service and what? What things were we gonna need to do? And that had to change quite rapidly as we learned more about it because none of us knew really.’’ S006*

The need for integration of care was quite evident from the beginning because different patients had different care needs, so different services needed to be activated or engaged with.

> *‘’when I did get into post, I was put in contact with respiratory consultant at the [redacted]… The COVID recovery lead. So, we what we did is near the beginning, you know when we started to see the patient, we started to work together a bit more and say well, let’s think about a pathway. … And I started doing that with cardiology. I started to do that with neurology. So, I started to create new pathways.’’ S001*

> *‘’we tried to connect them to other services which were already running within the trust, but also try to connect them to other clinicians within the system.’’ S006*

#### Knowledge

The new condition was still unknown to patients and professionals, each patient was different, and the array of symptoms was puzzling. The service provided an opportunity to develop and disseminate knowledge, which had a reassuring impact on some patients:

> *‘’ It was scary territory at the time because we did we we just didn’t know enough about it* …*we needed to find out more about it. The only way we could do that was by …developing these long COVID clinics that actually you can then monitor that person, post COVID*.’’ P005

> *‘’ And I think because I have knowledge, I wasn’t over worried about that*.’’ P009

> *‘’You know, no one told you this was what’s gonna happen. So you it’s kind of letting you know in advance. What could happen. I … I … I was quite happy to have that. I do feel that it’s needed*.’’ P011

The new condition was affecting patients in several ways, but the symptoms were all recognisable and issues staff members were used to treating. The uncertainty in the early days was soon replaced with the knowledge transfer from previous roles.

> *‘’I first was asked to join, it was very much like, you know, we don’t, we don’t really know what is out there…’’ S001*

> *‘’You, kind of like most clinical jobs that you do after you’ve been embedded into the ward or the service after a few months, it all comes …you have a confidence about what to do. But this job you don’t. Yeah, you just… It’s just 1/2 year later I still feel like a schoolgirl in this job.’’ S002*

Knowledge by the team, regarding the condition, the care needed and as shown to the patients, was a source of reassurance in a period when media messages created confusion.

> *‘’ There’s something about being able to reassure people in the context of the problems that they’re having, because we all built up our knowledge about some of the symptoms that people were having.’’ S003*

> *‘’I think we were at the start, we were slightly… You can’t know what you don’t know. … you were learning more about COVID, long COVID that then you’d think. … there was quite a lot of stuff that was coming out in the media about long COVID as well.’’ S006*

#### Reinforcement

Knowing about the service and feeling they were looked after, made an impact of patients’ wellbeing:

> *‘’ I think we just had a chat on the phone instead of went through all of my symptoms and umm, just kind of talked, talked it all through everything that was going on as far as I remember, which I found very reassuring… Umm, so I guess it made me feel, you know, hopeful*.’’P003

> *’’…hundred, 100% satisfied like I say. Just because it was such a new virus that we knew nothing about, it was just nice to know. That basically I’d had probably the best MOT in my life*.’’ P005

Positive feedback and patient outcomes reinforced the belief that staff members were heading in the right direction. Also, it was another source of reassurance for the patients that there was a way out of the uncertainty of their conditions. Reinforcing positive outcomes was also important to provide evidence to commissioners for more resources to be invested.

> *‘’You know that initial assessment and that review that I do is so thorough that they’re so they’re really delighted that someone has actually finally taken that whole story and is looking at that bigger picture.’’ S001*

> *‘’You still have to, have this, you know, kind of confidence of working in unknown uncertainty, but also the ability to kind of you know have confidence in like what we do know so far and what we have done so far has had a good effect.’’ S002*

> *‘’We had to move to a more generalist type approach with then specialist input… we kind of developed the offer based on what the patients were telling us.’’ S003*

## Discussion

### Statement of principal findings

The number of patients that accessed the long-COVID clinic during the first five months of implementation ranged between 15 and 30 per month, and patients were on average 5.75 months post initial COVID-19 infection when they were admitted. Patients attending the service were predominantly female, White British and were from non-deprived areas. The most commonly reported long-COVID symptoms reported by patients were shortness of breath and fatigue. On average there were 1.59 referrals out of the service to other specialists, with the majority of referrals going to pulmonary rehabilitation, primary care and community chronic fatigue specialists.

The emergence of a new virus with multiple post infective complications was one of the biggest health challenges in 2020. On a background of considerable uncertainty around pathology, optimum treatment and with very limited national guidance or research and in the face of increasing numbers of patients requiring support for LC symptoms, there was an immediate need to develop a new pathway in response to clinical need. Healthcare professionals within the NHS Trust had no preparation time or any opportunity to draw upon principles of established implementation or improvement science frameworks for creation of a pathway. Instead, staff acted rapidly according to patients’ needs and feedback and the resources available, as well as their previous experience, to meet patients’ needs.

The service went through a series of adaptations (i.e. implementation of data collection tools, inclusion of additional specialities) to deliver care based on the evolving understanding of the condition as well as patient’s needs. There was a substantial time lag between initial evidence of LC diagnosis and the ability to access care via the LC pathway, with some patients required to wait seven months for care. The delay in both the identification of LC as a distinct condition and acknowledgment of the need to invest resources for delivery of LC services at that time likely contributed to this. Nonetheless, the LC clinic delivered a high performing service that met patient needs during challenging times. When people entered the pathway, the journey was unpredictable and often included a reiteration of assessments and further referrals (S3 Figure). It demonstrates the cross-disciplinary collaborations that staff members established, or reinforced if already present, to provide high quality multidisciplinary care for complex LC care.

The establishment of a multidisciplinary team within the LC pathway, including a Specialist COVID-19 Rehab General Practitioner, Occupational Therapists, Physiotherapists, Speech and Language Therapists, Dietitians, Pulmonary Rehab, Chronic Fatigue specialists, and a Clinical Psychologist, enabled knowledge exchange across specialities. This contributed to increased knowledge of the condition, effective problem solving and the provision of a holistic approach to LC care.

Furthermore, due to their existing skills and capabilities HCPs were able to rapidly develop and establish further pathways into secondary care for patients that required more specialised investigations and review, which included onwards referral to respiratory, cardiology, ENT and neurology. Qualitative data confirmed the importance of the implementation of the new service for the patients’ physical and mental wellbeing. People felt looked after and in capable hands.

Professionals also felt positive about collaborative working, despite the challenging context. The pandemic gave HCPs the opportunity to work interactively with other services and deliver practical, integrated care pathways. HCPs expressed positive feelings around working in a multi-disciplinary team, collaborative working was perceived as satisfying and leadership was strong, staff members acknowledged the efforts and experience each person contributed to the creation of something new during exceptional times.

Patients and professionals, recalling the use and creation of the service, implied that the main areas to focus on, when implementing a new service for a novel condition, are the **capabilities of staff members**, specifically knowledge and expertise of health professionals delivering the service; **environmental context and resources**, to facilitate the initial implementation and ongoing delivery of a new care pathway; **knowledge** and prompt dissemination of information about the new condition, its consequences, and the new service; and positive **reinforcement**, when positive experiences and good patients outcomes creates reassurance and confidence in the way the system is adapted.

Overall, the experience of setting up a reactive and responsive service in an emergency was exceptional for all the people involved. Nevertheless, the already established organisational structure of the service supported the new pathway implementation well.

### Barriers and facilitators to care provided by the long-COVID pathway

Several barriers and facilitators to the implementation and delivery of the LC pathway, from the perspective of both patients and HCPs, were identified using the TDF [15]. See table 3. Barriers to the initial set-up of the LC pathway included budgetary constraints and the need to make a LC ‘business case’ to commissioners, lack of central face-to-face clinic location, lack of administrative and clinical processes and high workload. In addition, lack of knowledge around the disease characteristics of LC created uncertainty regarding which specialities were required within the pathway and multidisciplinary networks were created rapidly with limited prior planning. Difficulties setting up the service and subsequent access to this for patients may have contributed to the increase in average LC symptom severity in November 2020, as shown by the C19-YRS [27,28], although it is also possible that this reflected the timing of the second wave and lockdown, which created a barrier to access in-person facilities.

The present study has identified the value of transferable existing capabilities and expertise of health professionals, beyond their experience of specifically treating LC. In particular, the leadership experience from previous roles contributed to successful service implementation. Effective teamwork within the pathway and across different specialities was reported as a facilitator to both clinic set-up and delivery. The existence of a clinic tailored specifically to LC, and access to HCPs with specific knowledge of and expertise in treating the condition, provided reassurance for patients who were anxious and reported that they were “desperate”. HCPs understood patients needed dedicated time and acknowledgement of their condition, beyond that provided by usual care, and this served to validate patients’ experiences of LC. A finding which has been reported previously by patients receiving care in another multidisciplinary LC service [12]. In addition, patients reported excellent interactions with clinic staff and communication was reported as being good. Virtual delivery was well accepted by patients and perceived as a safe mode of delivery within this context. In contrast, virtual delivery presented some challenges for HCPs, including difficulties in completing necessary breathing assessments and a reduction in non-verbal cues. Nevertheless, virtual delivery did facilitate an efficient processing of heavy caseloads.

### Comparison with other studies

Current NICE Guidance recommends that people with LC should be offered integrated and coordinated care, referral for multidisciplinary assessment or, for specific complications, referral to specialist care [26], the opportunity to learn from expert by experience and professionals who managed the condition. Recent studies have reported on the implementation of support for LC within the UK health service context [14,33]. A recent prospective study of 1325 patients attending a dedicated LC assessment and management service set up in April 2020 [14], demonstrated the significant prolonged functional impairment of patients and the extensive need for onward referral to other specialists. A study that shows the potential barriers and facilitators of implementing such services will provide guidance for stakeholders.

Prior studies of UK LC clinics have identified several organisational and individual-level factors influencing their implementation and uptake, including the novelty of the condition, resource and capacity-related issues, knowledge limitations and lack of existing referral pathways [10,12].

However, to date there has been limited use of implementation science frameworks to explore this within this context. The Consolidated Framework for Implementation Research [34], a framework which can be used to identify contextual factors influencing successful implementation, was used to formulate interview questions for LC Leads in one recent study [12]. However, the framework wasn’t used to guide analysis and was not applied to the interviews with patients. In contrast to previous research the current study employed a mixed-methods research design which has facilitated a greater depth and breadth of understanding of the context within the LC service was implemented. Although the sociodemographic characteristics of those attending the LC service reflect that of the local population [35], patients from ethnic minority groups and areas of high deprivation were underrepresented, which reflects the findings of a previous study [14]. Patients from these groups were disproportionately affected by acute CoVID-19 infection [36,37] and arguably should therefore have been seen more frequently within LC services. This finding, however, may reflect lack of trust in health services, and experiences of inequalities of access of minority ethnic groups living with LC, a key barrier to uptake of services amongst this population [13].

### Strengths and weaknesses of the study

The current study is the first to our knowledge which includes a mixed methods analysis and has applied the TDF [15] to understand the barriers and facilitators to the implementation of a LC service. This led to evidence-based suggestions for the implementation of both future LC clinics and reactive services during critical public health emergencies. However, in the present study the TDF[15] was used retrospectively to guide analysis and was not used in the development of the qualitative topic guides, those were meant to let participants talk freely.

In addition, the study extends our knowledge of the lived experience of patients receiving care from multidisciplinary LC clinics as well as those involved in its delivery. The current paper reports on the implementation of one LC pathway, therefore it is likely that national and regional variations exist in the implementation and delivery of LC services within the UK health service context. As such, whilst four dominant TDF domains were identified as relevant to service implementation in the current study, there are likely to be other barriers and facilitators to the implementation of other LC pathways that haven’t been yet identified here.

In total, 102 people didn’t consent to share their data for analysis, which may have created bias in this respect. Moreover, of those eligible, very few patients agreed to participate in the interviews, which may be due to patient reluctance to recall difficult experiences related to LC or perceived additional burden at a time when they had competing demands in relation to managing their condition. Furthermore, interviews with patients and staff were conducted several months after the implementation of the service, contributing to the possibility of recall bias.

Quantitative data collection took time to be optimised when the LC clinic opened, the C19-YRS patient-reported outcome measure [27,28] was not yet available, and measures of anxiety and depression were initially only completed if there was perceived need to do so by clinic staff. A such, data for some measures were incomplete. However, adaptations to data collection processes later on during service delivery, further demonstrates the responsiveness of the service.

We were unable to collect data on the impact of the service on patients’ recovery, due to pragmatic reasons. A follow-up analysis is underway as part of another project. Despite these limitations, the triangulation of both the quantitative and qualitative data, alongside the inclusion of multiple perspectives, allowed for a deep understanding of the implementation of the LC pathway. This further increases the validity and reliability of the findings[38].

### Implications for clinicians and policymakers

The findings of the current study provide evidence-based recommendations for future practice in relation to the initial set-up, implementation, and ongoing delivery of new multidisciplinary LC clinics. Initial mapping of the ideal system of integrated care for long-COVID includes i) the identification of essential services based on patient needs’ assessment, ii) identification of existing and evolving knowledge of the condition amongst health professionals, and iii) the establishment of multidisciplinary referral pathways based on context and resources available. Furthermore, an integrated care approach, including clinicians from multiple disciplines, can be enhanced by the utilisation of implementation and improvement science frameworks [15,34] as part of the service development and planning. Collaboration with academic groups with expertise in implementation science and behaviour change can provide detailed insight into factors that are likely to impede or facilitate the successful delivery of service in context, prior to its implementation. Exploring the factors influencing integrated care processes and the mechanisms underpinning patient experiences of such processes, can support the promotion of patient-centred care [39]

The inclusion of multiple stakeholders in the planning process, including collecting data from health service managers, healthcare professionals and patients, can ensure the needs of all groups involved in the implementation and delivery of the service are met. Involvement of expert by experience (i.e. patients) in planning and management conversations, including providing the opportunity to express their needs to commissioning board meetings, can further ensure diversity and representation from underserved groups and mitigate risks to access.

Flexibility within the pathway is required, in particular it is recommended that contingency plans and adaptation routes are created in case the clinical condition evolves, and patients’ needs change. Additionally, identification of solutions to streamline the process (e.g., referral to multiple disciplines at the same time) is needed as soon as possible and, if the pathway seems to be effective, to consider the continuation of such process after the emergency has passed.

Finally, due to the requirement for multidisciplinary involvement and the need for streamlined inward and onward referral processes, the creation or adaptation of collaborative team working frameworks (e.g. communication strategies) should be embedded at the planning stage. Options for virtual interactions between patients and staff where appropriate, for instance beyond initial face to face assessments, should be explored, including homecare digital solutions. Provision of support for the wellbeing of staff members, including a physical space for breaks and opportunities for self-reflection and knowledge development can support staff wellbeing and consequently staff retention, protecting ongoing service delivery.

### Future research

There is a need for further qualitative research on patients’ lived experiences of receiving care from LC clinics, as well as an understanding of the longer-term impact on patient health and wellbeing of receiving multidisciplinary support for LC. Further research with patients discharged from the service whilst still symptomatic, of which there was a substantial proportion within the current study, would enable unmet need to be identified and would inform the provision of follow up care for these patients. Analysis of data collected from MDT LC clinics and related services across the UK is under way[40], to provide detailed information on the symptom and demographic characteristics and support needs of those attending. Collecting this data will provide a more complete picture of care for LC nationally and regionally, enable further knowledge exchange regarding effective rehabilitation for the condition, and provide evidence of gaps in access and reach of existing support. Research with patients from groups underrepresented in current research, and those overrepresented within the LC population, should focus on understanding the barriers and facilitators to accessing support to enable services to be tailored to their needs and thus increase uptake amongst this group. For this purpose, inclusion of data from primary and secondary care would be essential, to understand why some categories of patients were or not referred to services. Finally, we identified a potential theory of behavioural change that underpinned the evolution of services. An intervention could be developed using a template for process and quality improvement in community setting to evaluate its efficacy and for use in case of a public health emergency.

## Data Availability

All relevant data are within the manuscript and its Supporting Information files.

## Acknowledgements

The authors would like to thank Mr Mark Whiting, Dr Elizabeth Kendrick, Dr Robin Christie, Dr Jenna Chadwick, Dr David Wellsted for their support on this project, and Dr Jane Fry and Dr Alex Wilkinson for their helpful comments on earlier drafts of this manuscript. The authors would also like to thank the patients and healthcare professionals from the long-COVID service for their participation in this study.

## Supporting Information

**S1 Table. COVID rehab screening form**

**S2 File. Patient and HCP Interview topic guides**

**S3 Fig. Long-COVID pathway infographic flow diagram**

**S4 Text. Coding trees for patient and HCP interviews**

**S5 Table. COREQ checklist**

